# Human cytomegalovirus strain diversity and dynamics reveal the donor lung as a major contributor after transplantation

**DOI:** 10.1101/2022.03.02.22271774

**Authors:** Büsra Külekci, Stefan Schwarz, Nadja Brait, Nicole Perkmann-Nagele, Peter Jaksch, Konrad Hoetzenecker, Elisabeth Puchhammer-Stöckl, Irene Goerzer

## Abstract

Mixed human cytomegalovirus (HCMV) strain infections are frequent in lung transplant recipients (LTRs). To date, the influence of the donor (D) and recipient (R) HCMV-serostatus on intra-host HCMV strain composition and replication dynamics after transplantation is only poorly understood.

Here, we investigated ten pre-transplant lungs from HCMV-seropositive donors, and 163 sequential HCMV-DNA positive plasma and bronchoalveolar lavage samples from 50 LTRs with multiviremic episodes post-transplantation. The study cohort included D+R+ (38%), D+R− (36%), and D−R+ (26%) patients. All samples were subjected to quantitative genotyping by short amplicon deep sequencing, and 24 thereof were additionally PacBio long-read sequenced for genotype linkages.

We find that D+R+ patients show a significantly elevated intra-host strain diversity compared to D+R− and D−R+ patients (*P*=0.0089). Both D+ patient groups display significantly higher replication dynamics than D− patients (*P*=0.0061). Five out of ten pre-transplant donor lungs were HCMV-DNA positive, whereof in three multiple HCMV strains were detected, indicating that multi-strain transmission via lung transplantation is likely. Using long reads, we show that intra-host haplotypes can share distinctly linked genotypes, which limits overall intra-host diversity in mixed infections. Together, our findings demonstrate donor-derived strains as a main source for increased HCMV strain diversity and dynamics post-transplantation, while a relatively limited number of intra-host strains may facilitate rapid adaptation to changing environments in the host. These results foster targeted strategies to mitigate the potential transmission of the donor strain reservoir with the allograft.

## Introduction

Human cytomegalovirus (HCMV), a double-stranded DNA virus of the β-herpesvirus family, establishes a lifelong latent infection with reactivation episodes. Multiple HCMV strains (i.e. mixed infections) can be acquired during a person’s lifetime (Meyer-König et al., 1998)(Puchhammer-Stöckl et al., 2006) and are specifically frequent in transplant patients, where HCMV strains can be donor- or recipient-derived (D/R) or both (Puchhammer-Stöckl & Görzer, 2011). While infections in healthy adults are typically asymptomatic, they can lead to severe outcomes in those with immature or compromised immune systems (Fulkerson et al., 2021). Multiple strains provide an opportunity for viral recombination, selection for antiviral drug-resistant mutants and might consequently impact viral pathogenicity (Renzette et al., 2014). In fact, HCMV-infection related increased morbidity and mortality remains a high risk for lung transplant recipients (LTRs) (Almaghrabi et al., 2017).

With about 236 kbp in size, HCMV consists of predominantly conserved regions across strains and some highly polymorphic hotspots spread across the genome (Renzette et al., 2014). These variable loci result in a limited number of distinct genotypes that have been extensively used to study population-level HCMV diversity found within and between hosts, primarily focusing on glycoproteins such as gB, gN, gO and gH (Wang et al., 2021). Various techniques, including restriction fragment length polymorphism (RLFP) analysis (Huang et al., 1980), targeted amplicon sequencing (Puchhammer-Stöckl et al., 2006)(Coaquette et al., 2004)(Sowmya & Madhavan, 2009)(Hasing et al., 2021), and whole-genome sequencing (Dhingra et al., 2021)(Suárez et al., 2020) have been used. With increasing sequencing depth of next-generation sequencing platforms, the detection of low-frequency variants, i.e. minors became possible (Görzer et al., 2010). Currently, there is mounting evidence that HCMV exists as a heterogeneous collection of genomes with variations in composition and distribution between anatomical compartments (Renzette et al., 2013)(Hage et al., 2017) and over time (Dhingra et al., 2021)(Suárez et al., 2020)(Görzer et al., 2010)(Hage et al., 2017). However, in samples with mixed strains, determination of individual consensus sequences using short reads presents a challenge. In 2019, Cudini *et al*. introduced a computational method to reconstruct individual sequences within a sample from short-read data. They showed that the high nucleotide diversity of HCMV samples is due to mixed infections (Cudini et al., 2019). Another possibility to determine individual sequences in a sample is by single-read sequencing of long reads. We recently demonstrated that the true diversity in mixed populations of patient samples can be underestimated by short-read sequencing, since haplotype sequences sharing long stretches of sequence identity even in amplicon target regions can be missed, highlighting the utility of long reads (Brait et al., 2022).

Despite extensive research on HCMV strain diversity, much less is known about strain dynamics in LTRs, which can harbour heterogeneous viral populations (Görzer et al., 2008). In principle, dynamics can change by introducing new strains to the population, by reactivation of latent strains, *de novo* mutation, recombination or through the change in relative frequencies of present strains. It has been shown that reinfection with donor strains and reactivations of recipient strains can occur similarly often, although this is difficult to distinguish in mixed infected patients (Manuel, Pang, et al., 2009). Also, multiple strain transmissions from the donor organ to the recipient and a shift in strain predominance over time were found to be common (Hasing et al., 2021)(Hage et al., 2017), suggesting a complex dynamic post-transplantation.

Here, we combine Illumina deep-sequence and PacBio long-read sequence data from 163 specimens of 50 LTRs of different D/R risk groups with recurrent HCMV infection, to examine within-patient HCMV strain diversity and dynamics. This retrospective study points out major contributors to viral diversity in transplant patients leading to increased dynamics post-transplantation.

## Results

### PCR genotyping success rates of five genomic regions and overall genotype distribution

For this study, 50 LTRs with at least two HCMV DNAemia episodes with >10^2^ copies/mL either in the bronchoalveolar lavage (BAL) or EDTA-plasma (EP) or both, and one sample with >10^3^ copies/mL during the follow-up period of at least 185 days post-transplantation were included (details are provided in Materials and Methods). Genotypes of up to five polymorphic loci, namely UL6, UL73 (gN), UL74 (gO), UL139, UL146 (Figure 1) were assessed in 163 specimens consisting of 89 BAL and 74 EP samples by Illumina deep sequencing as described previously (Brait et al., 2022). This resulted in genotyping success rates for the five loci ranging between 67% and 85% (Figure 2A). Despite a comparable viral load distribution, BAL samples had a higher genotyping success rate for all loci than EP samples (Figure 2A). A detailed summary of PCR performances and the number of genotypes per locus are provided in Supplementary Table 1. We observed a trend towards higher viral loads with increasing numbers of maximally detected genotypes (Figure 2–figure supplement 1A). Considering the slightly different PCR genotyping success rates of the different loci, we also analysed each region separately (Figure 2–figure supplement 1B). Here, the association between higher viral loads and increasing genotype numbers was significant for gN (*P* = 0.0093), UL6 (*P* = 0.0490) and UL146 (*P* = 0.0031) but not for gO and UL139. The samples with the highest genotype numbers (≥3) did not have the highest viral loads. Of all samples with ≥ 2 genotypes at any locus 79% (49 out of 62) showed a single genotype in one of the five regions. These data indicate that the applied genotyping PCRs are highly sensitive also for samples with low viral loads starting from 10^2^ copies/mL, thus are well suitable for identifying mixed genotypes in clinical samples.

**Figure 1.**
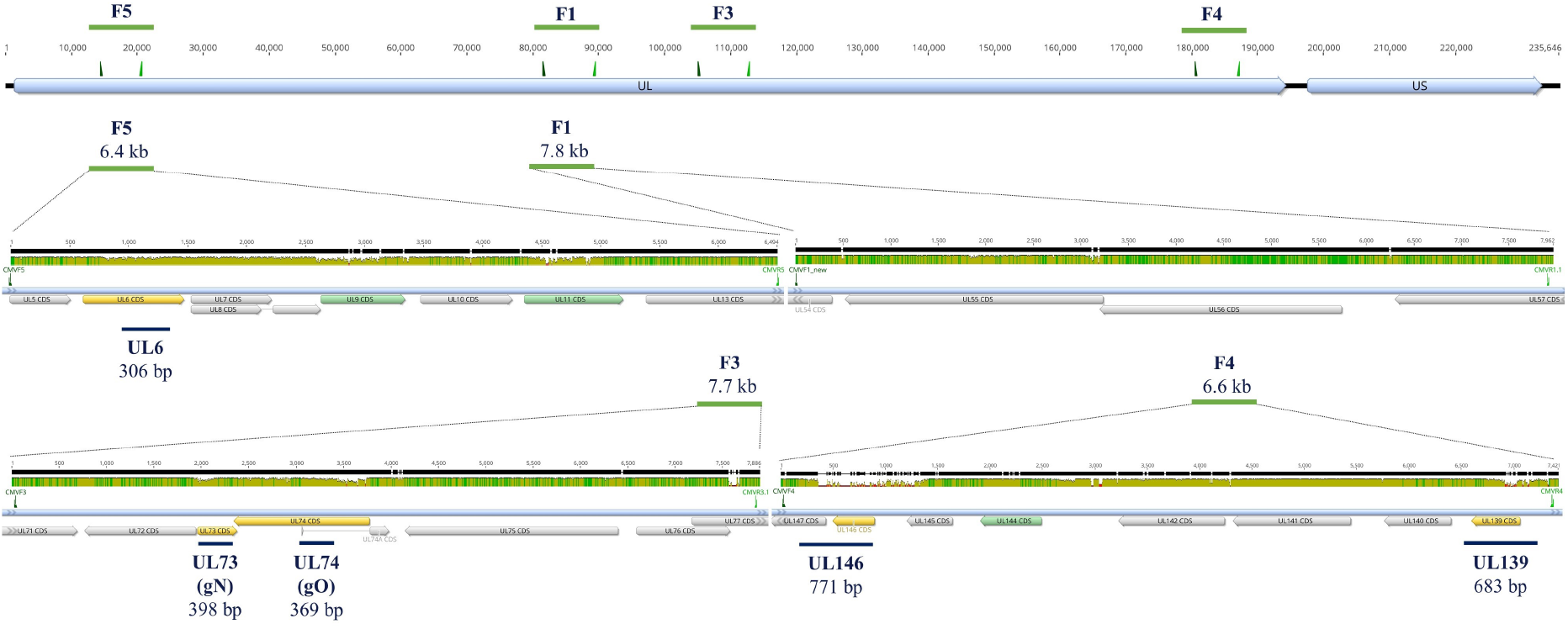
Locations of amplicon target regions along the HCMV genome of strain Merlin (NCBI: AY446894). The first track shows the overall HCMV genome structure that consists of a unique long (UL) and a unique short (US) region. Above, forward and reverse primers for the long amplicons (F5, F1, F3, F4) are depicted in dark and light green, respectively. Next, a zoom-in into these four long amplicons (green horizontal lines with amplicon sizes) are shown separately. The mean pairwise nucleotide identity for over 200 published sequences for these regions is shown in three colours: green (100%), brown (≥30% -<100%), red (≤30%). Annotated coding sequences (CDS) are shown in grey boxes and those used for short-read and long-read genotyping are orange and green, respectively. Lastly, blue lines below each long amplicon indicate short amplicon regions (UL6, UL73, UL74, UL146 and UL139) and the respective amplicon sizes. Graphs were generated with geneious prime 2019.0.3.

**Figure 2.**
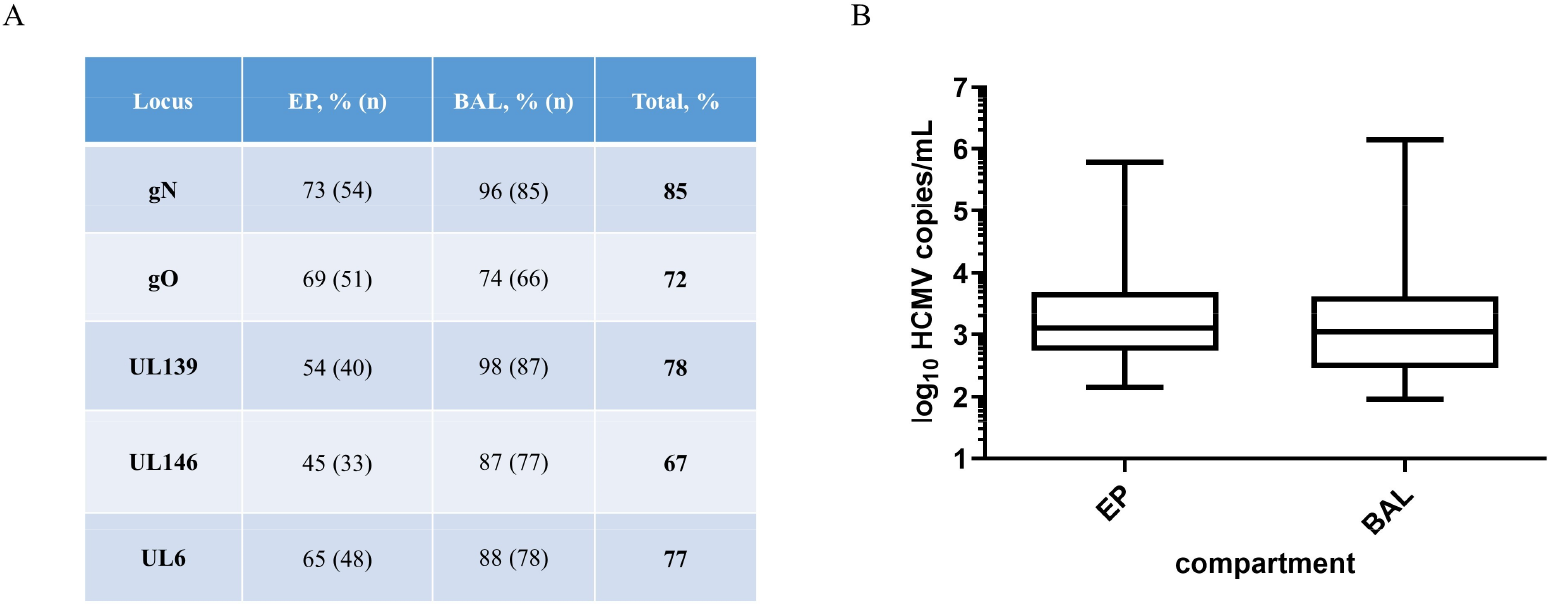
Genotyping performances for the five regions gN (UL73), gO (UL74), UL139, UL146 and UL6 and the viral load distribution for EP and BAL samples. **A:** Genotyping success in percent genotyped sample/total number of samples of this compartment. In total, gN has the highest PCR success rate and UL146 the lowest. **B:** Box whisker plots of the viral load of EP and BAL samples of the total cohort. The viral load distribution is not different between sample types (*P* = 0.301, n = 163, Mann-Whitney test). EP, EDTA-Plasma; BAL, bronchoalveolar lavage

We detected each genotype of the five loci in one and up to 20 LTRs, except for UL146-3, UL146-5 and UL146-6, which we did not find in any sample (Figure 2–figure supplement 2). Each genotype was also found as a major genotype (defined as >70% of all reads) in at least one sample. No significant differences between genotypes and viral loads were found for any of the five loci and this is illustrated for gO in Figure 2–figure supplement 3. These findings suggest that none of the detected genotypes had an obvious replication advantage over the others. The absence of UL146-3, UL146-5 and UL146-6, in contrast, may be indicative for functional differences among UL146 genotypes.

### Donor and recipient HCMV-seropositive patients display higher genotype diversity

We aimed to analyse the genotype diversity among the three HCMV serostatus combination groups, whereof 19 were D+R+ (38%), followed by 18 D-R+ (36%) and 13 D+R-LTR patients (26%). Similar HCMV DNA loads were observed between all three D/R serostatus combinations (Figure 3A). For each patient the maximum number of genotypes detected at any of the five loci and in any BAL and EP sample was counted (Figure 3B). More than one genotype was found in 84% of D+R+ patients compared to 62% and 50% in D+R- and D-R+ patients, respectively. D+R+ patients were significantly more frequently infected with > 2 genotypes (6/19) than D+R- and D-R+ patients (1/31) (*P* = 0.0089; Fisher’s exact test). Moreover, the D+R+ patient group shows the highest numbers of genotypes with up to four genotypes in two LTRs (Figure 3B). These data suggest that both, lung donor and recipient contribute to the HCMV strain diversity in D+R+ patients.

**Figure 3.**
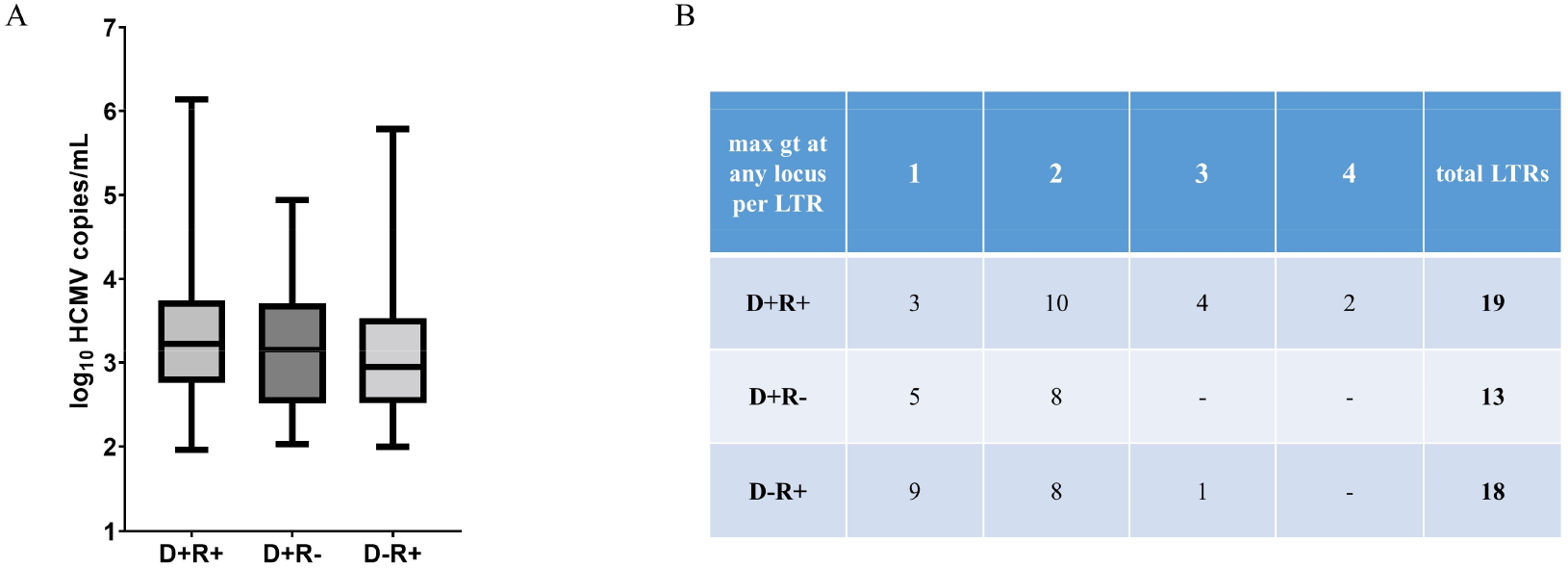
Viral load and maximum number of genotypes (max gt) in lung transplant recipients (LTRs) with different HCMV serostatus combinations of donor (D) and recipient (R). **A:** No significant difference in viral load between the three HCMV serostatus combination groups is observed (*P* = 0.2626, n = 163, Kruskal-Wallis test plus *post hoc* Dunn’s multiple comparisons test) **B:** The maximum number of genotypes (gt) at any of the five loci detected for each LTR is provided (n = 50).

### Donor HCMV-seropositive patients exhibit higher genotype dynamics over time

Next, we analysed how the genotype composition within a patient changes over time in the three serostatus patient groups. In 36/50 LTRs genotyping data of at least two episodes of HCMV DNAemia in the same compartment were available and allowed analysis of intra-host genotype dynamics over time (Figure 4A). In total, 26 BAL and 14 EP sample pairs were analysed (Supplementary Table 2). The median time difference between the paired samples was 179 days (range = 14 - 531 days). LTRs with at least one sample with ≥ 2 genotypes at any loci or differing genotypes at different time points were defined as mixed infected (n = 27). On the contrary, in single infected patients one genotype was identified longitudinally (n = 9). We defined the total genotype dynamics over time as a change in two variables: 1) increase in genotype number and 2) change in relative genotype frequency. The former describes an increase in genotype number between time points 1 and 2, while the latter accounts for a predominance change in the major genotype (defined as >70% of all reads) between the two episodes.

**Figure 4.**
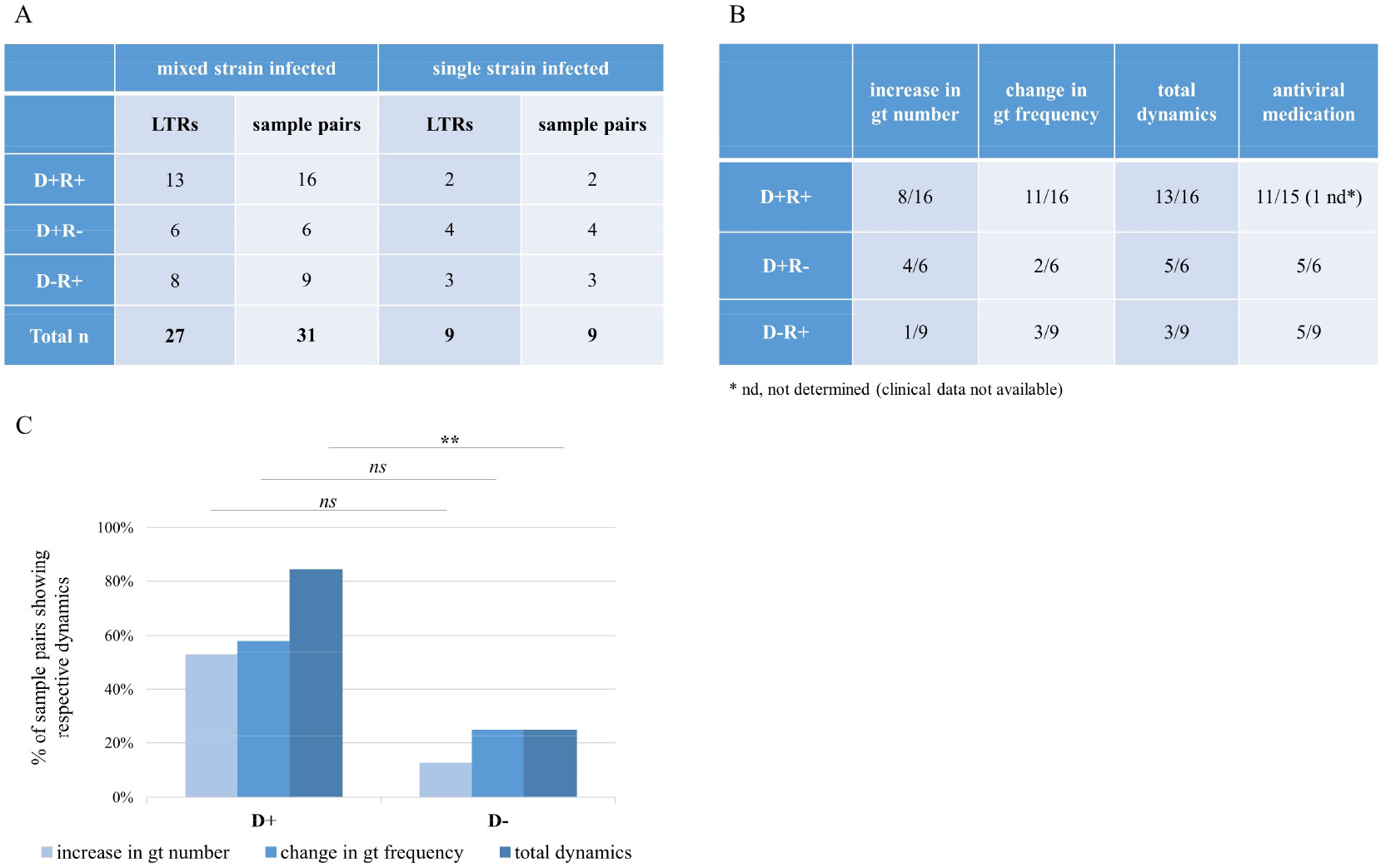
Intra-host genotype dynamics over time in lung transplant recipients (LTRs) of the three HCMV-serostatus combination groups. **A:** The numbers of LTRs and the number of sample pairs comprising the dynamics cohort are categorised into the three serostatus groups. LTRs from whom samples from two pairs were available, both were analysed. **B:** The table shows the number of mixed sample pairs that presented the respective dynamics, increase in gt number, or change in gt frequency, or both (total dynamics). Antiviral medication was considered when given between or at time of analysed time points as a fraction of all pairs in this group. **C:** For statistical tests, one sample pair per patient (the earliest one tested) was included (n = 27, Fisher’s exact test; ** for *P* < 0.01). Total dynamics was significantly higher in D+ patients compared to D-patients (*P =* 0.0061), while the increase in gt number and change in gt frequency did not reach significance (*P =* 0.0899 and *P* = 0.2087; respectively). gt, genotype; D, donor; R, recipient; ns, non-significant

D+ patients showed significantly higher total dynamics than D- patients (*P* = 0.0061) (Figure 4B, 4C) and a trend in the increase in genotype numbers (*P* = 0.0899). In 69% of D+R+ patients a change in predominance was observed compared to 33% each in the other two patient groups. Antiviral medication, which may influence strain dynamics, was similarly frequent between single- and mixed-infected LTRs (*P* > 0.9999, n = 34), between mixed-infected D+ and D-LTRs (*P* = 0.1972, n = 26), and between LTRs with and without total dynamics (*P* > 0.9999, n = 26). These data indicate that there is no significant medication effect on strain dynamics.

### Two patterns of genotype predominance dynamics over time

The predominance change in relative genotype frequency displayed two distinct patterns. First, an exchange of the pre-existing genotype (thereby an initially minor genotype becomes the major), and second, the introduction of a new genotype that overtakes the previously predominant genotype (whereby the initially major genotype becomes undetectable or a minor). While paired BAL samples (n = 10) presented both predominance patterns equally frequent, all plasma sample pairs (n = 6) displayed the second pattern, introduction of a new predominant genotype (Supplementary Table 2). Overall, a change in predominance was detected the earliest 62 days after the first episode, indicating that intra-host dynamics can be rapid.

### The pre-transplant donor lung may harbour multiple genotypes

To learn more about the donor lung as a potential HCMV source, we analysed the middle lobe part of ten lungs of HCMV-seropositive donors for HCMV-DNA positivity and genotype diversity. Of each middle lobe, one to eight different locations were collected (Supplementary Table 3a). Each individual tissue piece was stored in buffer before being processed to a single cell suspension (details see Materials and Methods). In total, 60 lung pieces and the corresponding storage buffer samples were tested for HCMV positivity. We detected HCMV-DNA in 8/60 cell suspensions and in 15/60 storage buffer samples. Frequent detection of HCMV-DNA in storage buffer is most likely due to release of HCMV-DNA from damaged cells and/or blood vessels during collection. In summary, five out of the ten lung donors were HCMV-DNA positive. Two out of ten donors were HCMV-DNA positive in the cells only, two donors were positive in the storage buffer only and one donor was positive in both, cells and storage buffer. In the cellular fractions, we detected HCMV-DNA in up to four out of eight different locations (Supplementary Table 3a). These findings indicate focal HCMV-DNA distribution in the analysed lung tissues. All HCMV-DNA positive samples were subjected to short amplicon genotyping. We detected up to three genotypes in a single donor sample and three out of four donors displayed mixed genotypes (Supplementary Table 3b). These findings show that multiple HCMV strains are transmittable with the donor lung, thus may substantially contribute to an increased HCMV strain diversity post-transplantation.

### Long-read PacBio sequencing for haplotype analysis

Thus far, we have assessed HCMV strain diversity and dynamics based on sequencing data of short polymorphic regions to define the respective HCMV genotypes. To expand our understanding on within-host HCMV strain diversity we quantitatively determined the individual haplotypes in a subset of 23 BAL and one EP sample from 20 LTRs by long-read PacBio sequencing. Sample selection was restricted by the viral load sensitivity limit of the long-read PCR (for details refer to Materials and Methods). Herein, the term haplotype refers to a single HCMV genome of > 6kb covering multiple non-adjacent genotype-defining regions.

Long amplicons were generated from four regions, F5, F1, F3 and F4 (Figure 1) and subjected to long-read PacBio sequencing similarly as described previously (Brait et al., 2022). The amplicon fragments range between 6.2 and 7.9 kb in length and cover all polymorphic genes (dN, nonsynonymous substitutions per nonsynonymous site ≥ 0.086) we used for genotyping and 15 more. These include seven of the most divergent HCMV genes (dN ≥ 0.034) and two less variable genes UL55 (gB) and UL75 (gH), extensively sequenced in previous studies due to their known functional importance (Sijmons et al., 2015). Detailed information on PacBio-determined haplotypes are presented in Supplementary Table 4a. In Figure 5A the assignments of the identified haplotype sequences to the most similar reference sequences are displayed. Ten LTRs were single haplotype infected, nine LTRs were infected with up to two haplotypes, and in one patient (LTR-45) up to three haplotypes were detected. In total, we identified 116 individual haplotypes in 20 LTRs and for each, sequences with identities >98% were found in the NCBI Nucleotide (nr/nt) database (Supplementary Table 4b), except for four haplotypes sharing only 93% to 96% identity. Notably, for 38% of the 165 reference sequences corresponding haplotype sequences were found in our cohort illustrating a substantial inter-host haplotype diversity. Despite a high inter-host variability, we found haplotypes in different patients sharing sequence identities of up to 99.9% and 99.8% in F3 and F5, respectively (6A_hap1, 12B/F_hap1) and 99.7% in F1 (16B_hap2, 45B/D_hap2). Haplotypes of F4 were more diverse with a maximum of 98.5% sequence identity between haplotypes 11O_hap1 and 37B_hap1.

**Figure 5.**
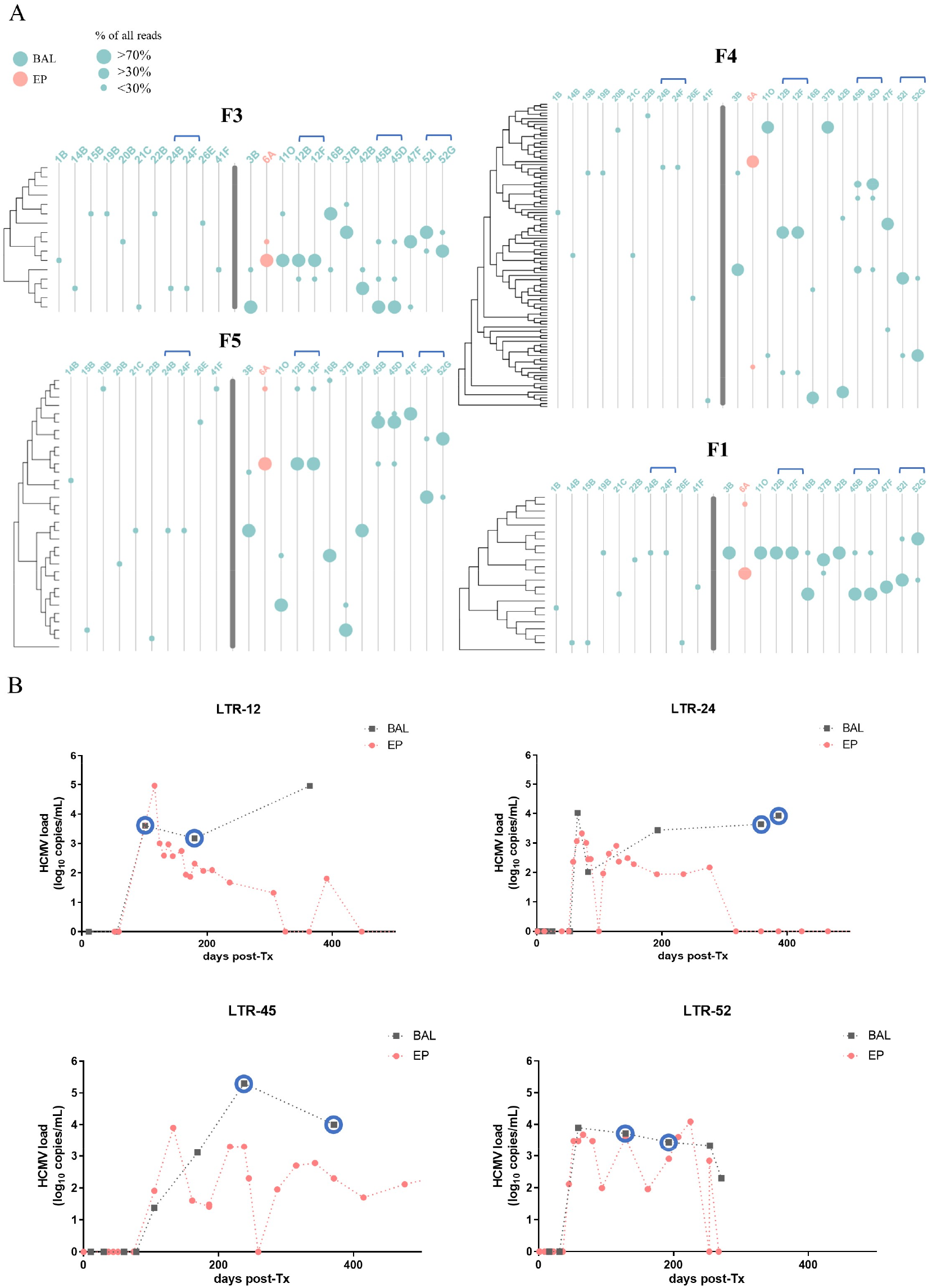
**A: Graphical representation of the 144 detected haplotypes of the four long amplicon regions.** The trees cluster representative haplotypes of the respective regions that differ in >150 nucleotides based on all publicly available HCMV strains (accession numbers are provided in Supplementary Table 7a). Distances are in the units of the number of base differences per sequence. Trees were generated in MEGA X using the UPGMA method and displayed with Evolview. Single and mixed infected samples are separated by a bold grey line. Samples 24B/F,12B/F, 45B/D and 52I/G are follow-up samples of the same patients and are indicated with blue brackets. **B: HCMV viral loads of longitudinal samples from BAL and EP from LTRs 12, 24, 45 and 52**. Blue circles indicate samples that have been subjected to PacBio long-read sequencing. BAL, bronchoalveolar lavage; EP, EDTA-Plasma; LTR, lung transplant recipient; post-Tx, post-transplantation

### Rapid intra-host dynamics, but stable haplotype sequences over time

For four LTRs two follow-up BAL samples each were included (Fig. 5A; indicated with blue brackets) to assess the haplotype dynamics over time. The course of HCMV-DNAemia for these four patients is provided in Figure 5B. First, one patient (LTR-24) was single haplotype infected showing the same haplotypes in both samples (24B, 24F; time between samples (Δ): 28 days). Second, the longitudinal samples 52I and 52G (Δ: 64 days) of LTR-52 illustrate the change in predominance of the two haplotypes from time points 1 to 2 throughout all four regions. Third, samples 12F and 12B (Δ:79 days) of LTR-12 show the same two haplotypes with consistent relative frequencies for both time points. Fourth, for patient LTR-45 a change in predominance of the major haplotype is only observed in region F4 (Δ:133 days). Alignments of the within-patient haplotype sequences show 100% sequence identity except for the minor haplotypes of samples 45B and 45D for the region F1 (45B/D_F1_hap2) and a suspected fourth haplotype for region F4 (45B_F4_hap4). Here, a closer look reveals that 45B/D_F1_hap2 consists of a mixture of two sub-haplotypes with 12 nucleotide differences (Supplementary Table 4c). Separation of the two sub-haplotype sequences show that both sub-haplotypes are present in samples 45B and 45D and the respective sequences are 100% identical in both samples. The sequence of haplotype 45B_F4_hap4, in contrast, is different. Visual inspection of the individual PacBio circular consensus sequence (ccs) reads revealed two different groups of ccs, both sharing sequence similarity with haplotype sequences 45B_F4_hap1 and 45B_F4_hap2 (Figure 6–figure supplement 1). The two groups with 12 and 15 ccs reads each display recombined sequences with breakpoints upstream and downstream of coding sequence (CDS) of UL144, respectively. PCR-mediated recombination cannot be ruled out, thus, these sequences were excluded from further analysis. In summary, the longitudinal haplotype data show, as observed for the genotyping data, that changes in predominance can be fast-paced, while the haplotype sequences remain stable.

### Genotypes shared between intra-host haplotypes limit overall diversity in mixed samples

To investigate how linkage combinations between non-adjacent polymorphic regions contribute to intra-host diversity in mixed strain samples, we compared the number of genotypes of eight highly polymorphic genes (dN≥0.082) with the number of haplotypes in these regions. In total, 10 mixed infected LTRs with 75 distinct haplotypes were analysed. Genes used for genotype determination were gN and gO in F3, UL139, UL144 and UL146 in F4 and UL6, UL9 and UL11 in F5 (Figure 1). For region F3, in all samples the number of genotypes matched the number of haplotypes, possibly because of the well-described linkages between gN and gO genotypes restricting recombination and consequently different linkage combinations (Mattick et al., 2004). All haplotypes in our cohort showed previously described gN/gO linkages. In three LTRs fewer genotypes than haplotypes were observed in target regions F4 and F5, suggesting different linkage patterns (Supplementary Table 5). First, we identified two F5 haplotypes in sample 3B, both sharing the UL11-2 genotype with only one nucleotide difference in the 429 bp long region used for UL11 genotyping. Pairwise alignments of the two F5 haplotypes clearly show substantial differences across the full 6.4 kb sequence with stretches of high identity in UL11 CDS (Figure 6–figure supplement 2A). Second, we found two F4 haplotypes in sample 11O, but only one UL139-2 genotype with 14 nucleotide differences (98% identity) across the genotyping region (Figure 6–figure supplement 2B). Lastly, in both samples of LTR-45 (45B/45D) three F4 haplotypes, but two UL139, UL144 and UL146 genotypes and three F5 haplotypes, but two UL6, UL-9 and UL11 genotypes were found. Again, haplotypes differed in one and up to 11 nucleotides in the respective genotyping regions, but considerable differences up-and downstream confirmed distinct haplotypes (Figure 6–figure supplement 2C and 2D). Interestingly, while the same UL139 genotype was shared between hap1 and hap2, the same UL144 and UL146 genotypes were found between hap2 and hap3, which is indicative of past recombination. Of note, intra-host haplotypes in our cohort shared up to 40% identical sequence sections, which is ideal for homologous recombination (Figure 6-figure supplement 3). Taken together, the full haplotype sequence allowed a clear determination of distinct haplotypes, although same genotype assignments were found.

While linkage patterns of non-adjacent genotypes within a haplotype region can unambiguously be defined by long read sequencing, linkages of non-overlapping haplotypes might be assessed according to their relative frequencies (Brait et al., 2022). We refer to this as a statistical linkage of haplotypes (relative frequencies of all haplotypes are provided in Supplementary Table 4a). As illustrated in Figure 6B for patient LTR-52 it can be assumed that the major (hap1) and minor (hap2) haplotypes, respectively, are present on the same HCMV genome. Moreover, the statistical linkage remained stable even after the predominance change from time point 1 to 2. For samples with varying relative frequencies of the distinct haplotypes of each region, direct statistical linking is not reasonable. The longitudinal samples 45B/45D shown in Figure 6A, exemplify this problem clearly. First, in both samples the relative haplotype frequencies were similar for the regions F5, F1, and F3, but not for F4. Second, the major haplotype of F5, F1, and F3 did not change over time, but the major haplotype of F4 did. Third, relative frequencies of the haplotypes, hap2 and hap3 were only similar for F5, F1, and F3, but not for F4. Thus, the three individual haplotypes of F5, F1, and F3 can be statistically linked, but no further linkage with the F4 haplotypes is feasible. These data may indicate that more than three unique HCMV strains are present in these samples, probably resulting from recombination in between the target regions F3 and F4.

**Figure 6.**
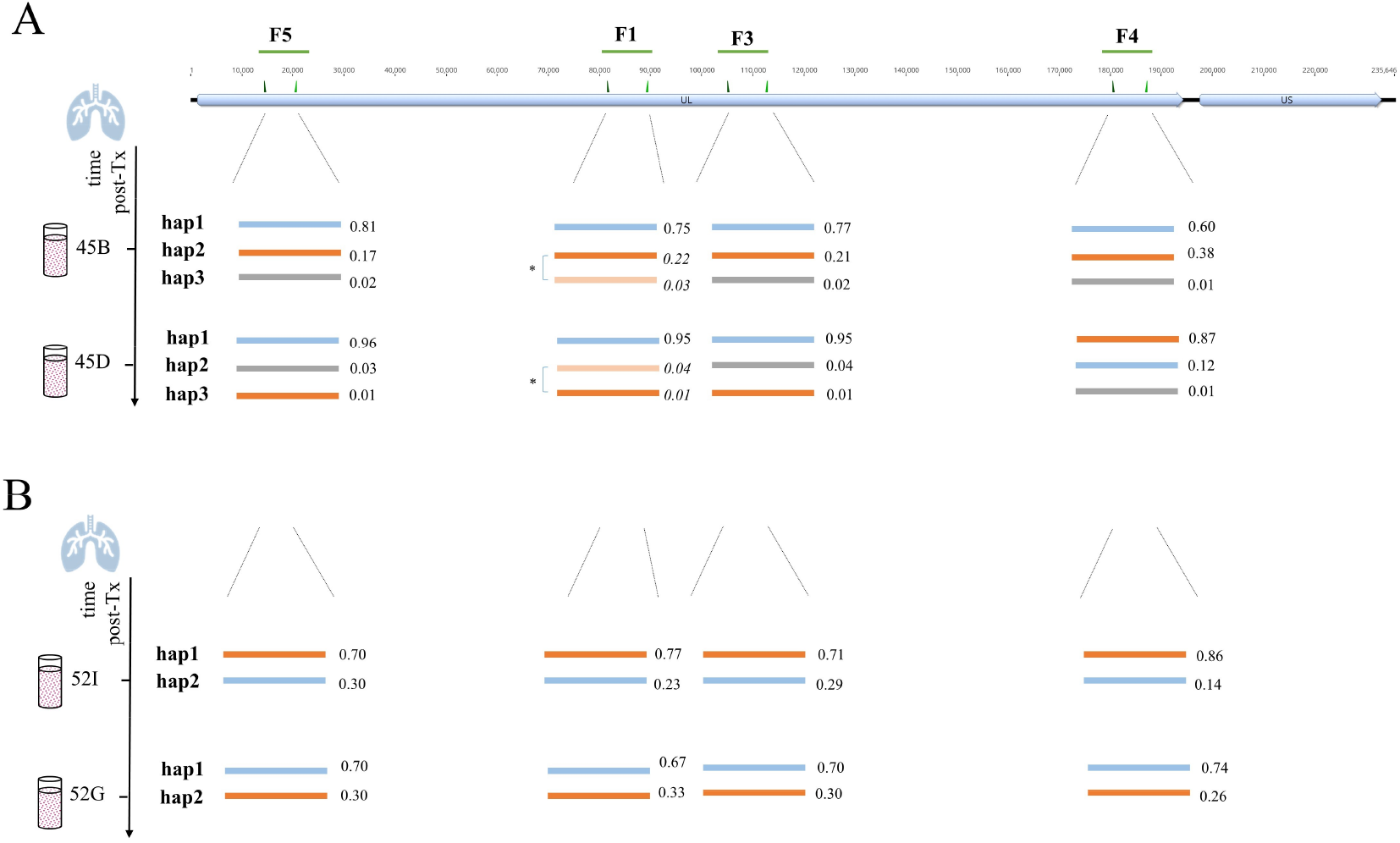
Schematic presentation of haplotypes of longitudinally PacBio sequenced BAL samples of LTR-45 (A) and LTR-52 (B). The top track shows the overall HCMV genome structure that consists of a unique long (UL) and a unique short (US) region with the long amplicon target regions (F5, F1, F3, F4) depicted as green lines. Haplotypes determined for each of the four long amplicon regions are presented as lines of different colours. The values next to each haplotype are the relative frequencies of the respective haplotype. **A:** The two sub-haplotypes of LTR-45 of region F1 (marked with an asterisk) were initially detected as a single haplotype (hap2), since they only differed in 12 nucleotides across the whole fragment (further details in Results and Supplementary Table 4c). Here, there are shown as separate haplotypes and the calculated relative frequencies are shown in italic. **B:** For LTR-52, similar relative frequencies of both haplotypes of each region suggest their statistical linkage. A change in predominance is observed between both time points and support that orange and blue haplotypes are linked, respectively. post-Tx, post-transplantation; nt, nucleotide

## Supplementary Figures

**Figure 2–figure supplement 1.**
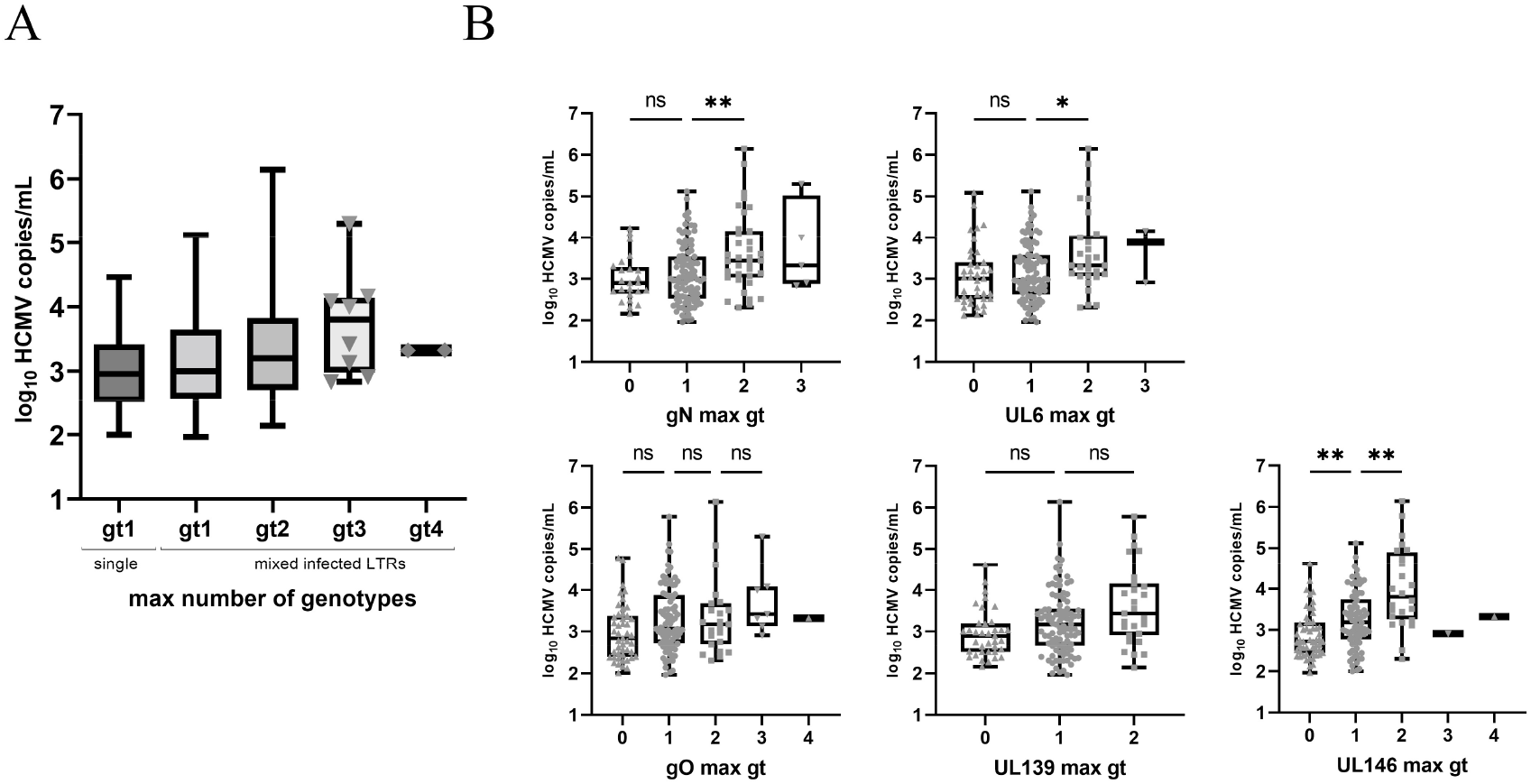
Box and whisker plots of the HCMV viral load (VL) and the maximum number of genotypes (max gt) detected for all five loci or for gN (UL73), gO (UL74), UL139, UL146 and UL6 separately. **A:** A trend towards higher VL with increasing gt numbers is observed, but is not significant (*P* = 0.0858, n = 161). Symbols show each sample point for groups, where the total n was < 40. Single and mixed LTRs are shown separately. In the single group are samples with a single gt at each locus, while samples in group gt1 (mixed infected LTRs) had only one detectable gt in this sample but belong to a mixed infected patient. **B:** All sample points are shown as grey symbols. Max gt 0 shows the VL of PCR-negative samples. VL are significantly higher for samples with max gt = 2 compared with gt = 1, for gN (*P* = 0.0093, n = 134) and UL6 (*P* = 0.0490, n = 122). For UL146 PCR, samples with max gt = 0 compared with max gt = 1 and max gt = 1 compared with max gt = 2 are also significantly different in their VL (*P* = 0.0075, n = 137 and *P* = 0.0031, n = 106, respectively). Kruskal-Wallis test with *post hoc* Dunn’s multiple comparison test was applied for samples with n > 5 (* for P < 0.05; ** for P < 0.01) ns, non significant; LTR, lung transplant recipient; gt, genotype

**Figure 2–figure supplement 2.**
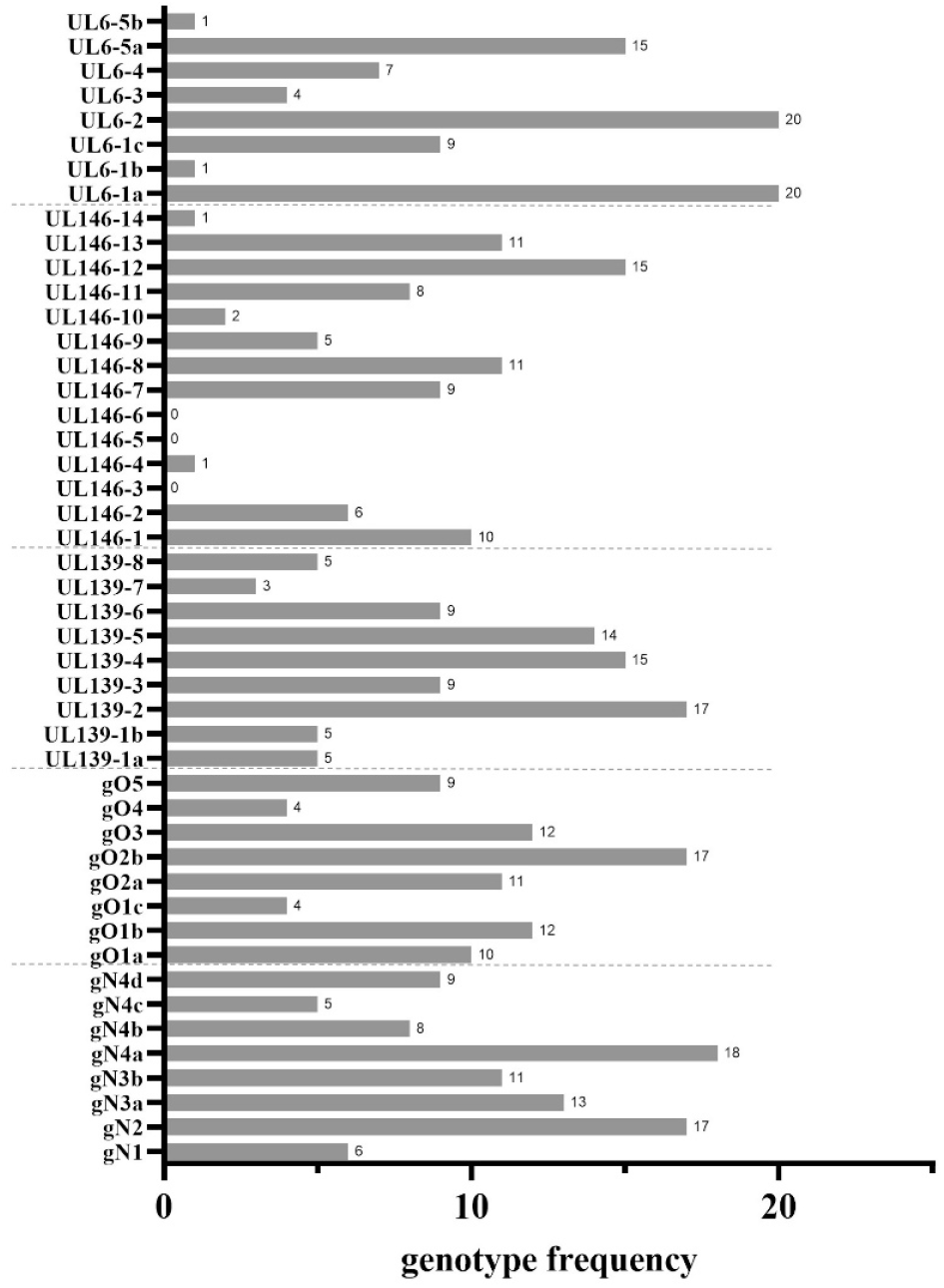
Normalised frequencies of all genotypes of five polymorphic loci in 50 lung transplant recipients (LTRs). If a certain genotype occurred multiple times in different samples of the same LTR, it was counted once for this analysis (normalised frequency). Both, plasma and bronchoalveolar lavage samples are included. We find all genotypes except for UL146-3, UL146-4 and UL146-6.

**Figure 2–figure supplement 3.**
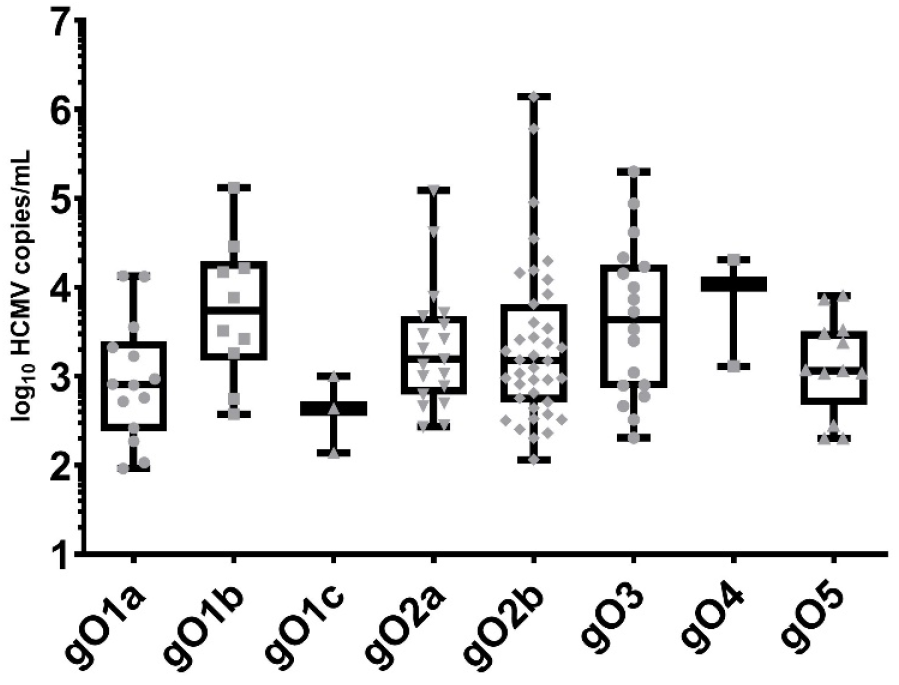
Major gO genotypes detected in 50 lung transplant recipients. Only major genotypes (defined as >70% of all reads) are shown and used for analysis, since the major strain is expected to contribute the most to the total viral load. No significant difference between viral loads and genotypes were found in a Kruskal-Wallis test with *post hoc* Dunn’s multiple comparisons test for groups with n > 5 (*P* = 0.1189, n = 112).

**Figure 6–figure supplement 1.**
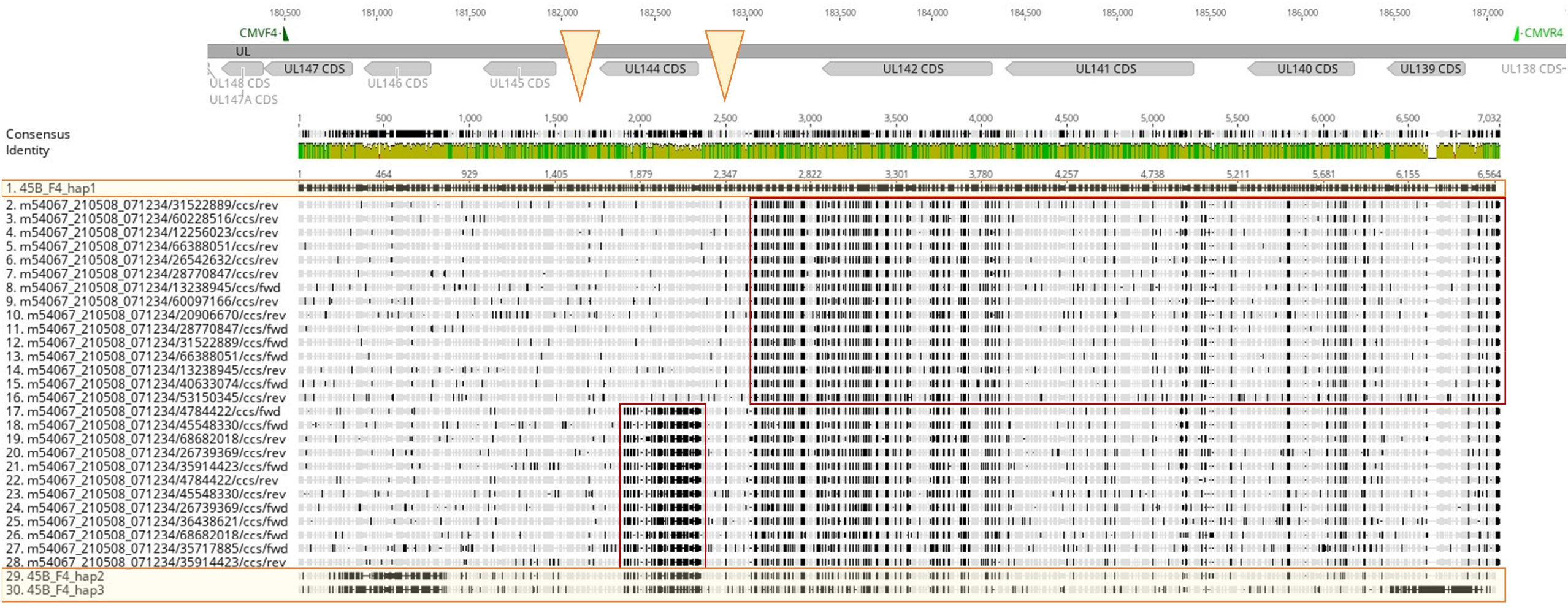
Alignment of the three haplotypes of sample 45B of region F4 (45B_F4_hap1, 45B_ F4_hap2 and 45B_ F4_hap3) with the PacBio circular consensus reads (ccs) that were initially aligned to the fourth haplotype 45B_F4_hap4. The upper track displays the coding sequences (CDS) along the 6.6 kb F4 amplicon region, below the mean pairwise identities of the aligned sequences are shown in green (100%), brown (≥30% -<100%) and red (≤30%). Orange triangles indicate potential breakpoint regions for recombination. Then, the alignment of 30 sequences is shown with hap1 set as reference (top sequence marked in orange). The sequences between the orange highlighted haplotypes at the very top (hap1) and bottom (hap2 and hap3) are the 27 ccs reads belonging to hap4 in the first round of mapping. This alignment illustrates that the haplotype sequences that were used to construct the consensus sequence of hap4 actually consists of two groups and indicate two regions of recombination. The upper 15 ccs reads show similarities to hap1 before switching to hap2 around the position of the 2^nd^ triangle. The other 12 reads switch at the 1^st^ triangle. Red boxes show where the change into the pattern of hap2 occurs.

**Figure 6–figure supplement 2.**
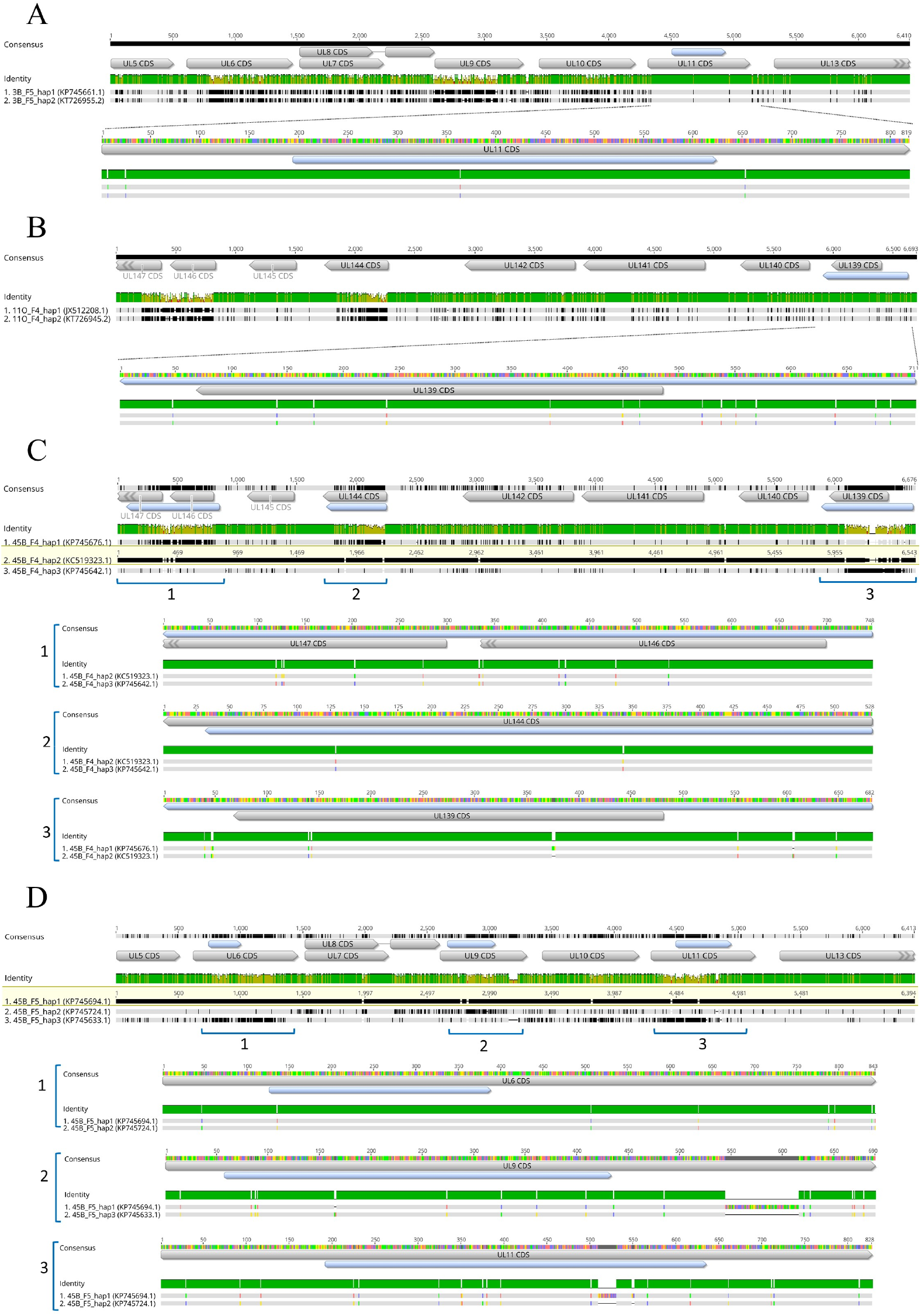
Alignments of intra-host haplotypes of samples 3B (A), 11O (B), 45B F4 (C) and F5 (D) show minor differences in genotype-defining regions. In the upper track the coding sequences (CDS) are displayed in gray boxes. The regions used for genotype determination are shown in light blue. Below, the mean pairwise identities of the aligned sequences are shown in green (100%), brown (≥30% -<100%) and red (≤30%). Next, intra-host haplotypes are aligned and differences shown with black lines, with one haplotype set as reference in C and D (highlighted in yellow). In the zoom-in sections below, the two haplotypes sharing genotypes in the respective region are pairwise aligned and the nucleotide differences are shown in colours corresponding to the respective nucleotide (green, thymine; blue, cytosine; red, adenine; yellow, guanine).

**Figure 6–figure supplement 3.**
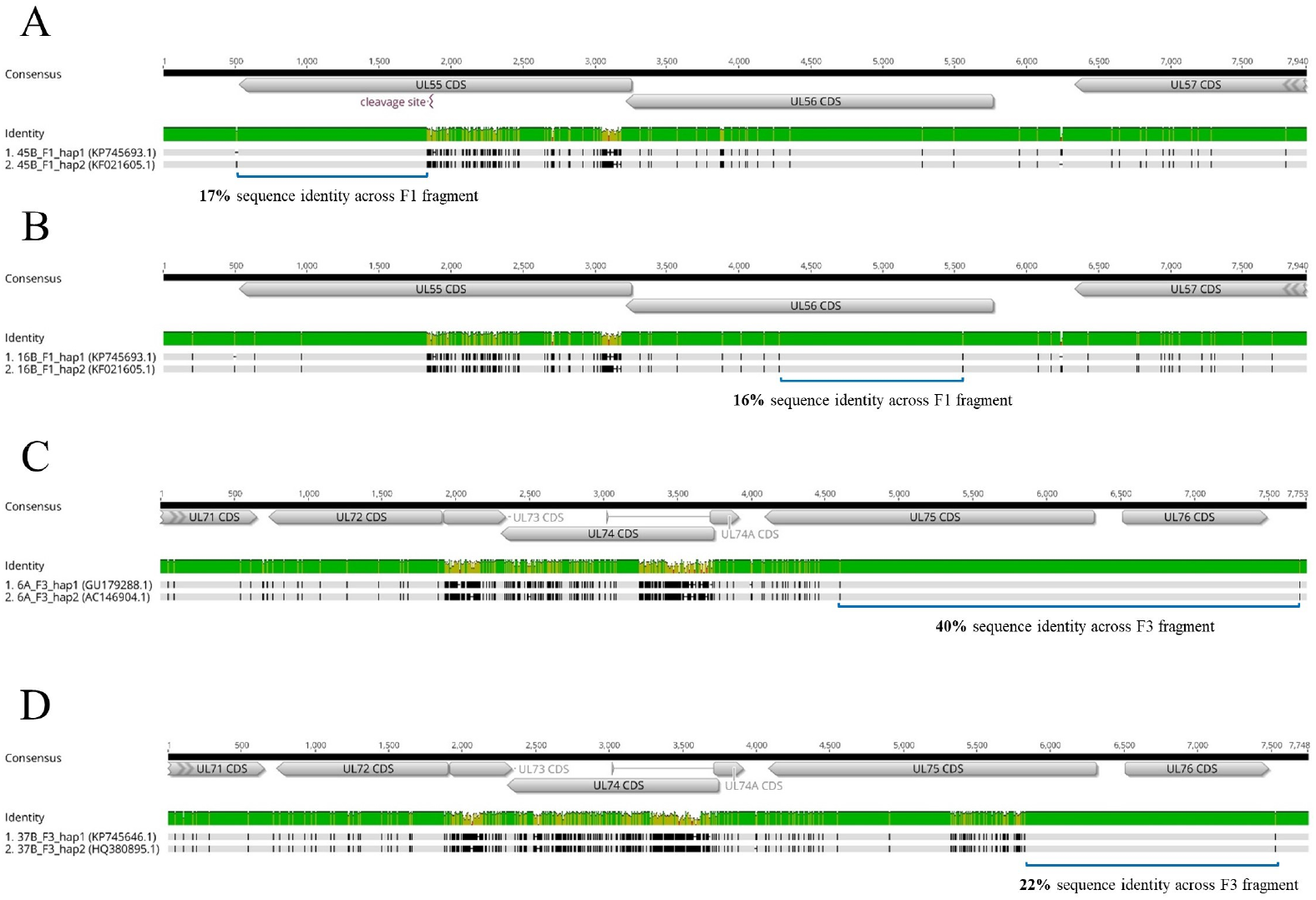
Sequence alignments of intra-host haplotypes of samples 45B, 16B, 6A and 37B show long stretches of sequence identity. In the upper track the coding sequences (CDS) are displayed in gray boxes. The regions used for genotyping are shown in light blue and the cleavage site in gB is marked. Below, the mean pairwise identities of the aligned sequences are shown in green (100%), brown (≥30% -<100%) and red (≤30%). Lastly, intra-host haplotypes are pairwise aligned and differences shown with black lines.

## Discussion

In this study, we have comprehensively assessed the diversity and replication dynamics of mixed HCMV strains in patients with multiviremic HCMV episodes after lung transplantation. We identify the donor lung as a critical source for a complex and dynamic HCMV strain population independent of the recipients’ HCMV serostatus and we demonstrate that distinct genotype linkage combinations may not necessarily increase the overall intra-host diversity.

Sensitive and quantitative assessment of HCMV genome diversity directly from clinical material is important to study the composition and dynamics of mixed infections robustly. Due to the low amount of starting HCMV-DNA in clinical specimens, PCR-amplicon enrichment followed by short-read Illumina deep-sequencing was performed. Previous studies using PCR-based approaches confirmed that the detection of mixed strains could depend on the viral load, the genetic loci and clinical specimen type analysed (Görzer et al., 2008)(Sowmya & Madhavan, 2009)(Manuel, Åsberg, et al., 2009). We were able to analyse samples with viral loads starting from 10^2^ copies/mL and detect minor variants down to 1%, making this approach explicitly more sensitive for detection of mixed infections compared to a whole genome sequencing approach (Suárez, Wilkie, et al., 2019). The five genes (UL6, gN, gO, UL139 and UL146) we chose are among the 12 highest polymorphic ones with a dN ≥ 0.086, are predicted to encode for products of immunomodulation and are known to be necessary for cell entry (Sijmons et al., 2015)(Wang et al., 2021)(Bradley et al., 2008). While the high overall genotyping success rates (67-85%) confirm that these target regions are very well suitable to determine genotypic diversity the data also show that HCMV-DNA isolated from BAL samples is better amplifiable than from plasma samples (Figure 2A). This is most likely due to the high fragmentation of plasma HCMV-DNA as previously suggested (Tong et al., 2017)(Brait et al., 2022). This also explains why amplicons of > 6kb, which were used for long read sequencing, could hardly been generated from plasma-derived HCMV-DNA, despite similar viral load concentrations as in BAL samples.

In our cohort of 50 lung transplant patients, we found representatives of almost all genotype sequences (41 out of 44), yet with varying prevalences. No specific genotype was associated with significantly higher or lower viral loads, suggesting that genotypic variations do not affect HCMV replication efficiency (Figure 2–supplement 2). Three UL146 genotypes, gt3, gt5 and gt6, were not present in our study cohort (Figure 2–supplement 3). This is well in accordance with prior studies also reporting a low frequency for these genotypes (Bradley et al., 2008)(Berg et al., 2021). UL146 ORF encodes a viral α (CXC)-chemokine, that is suggested to recruit neutrophils for viral dissemination (Lüttichau, 2010). A recent study investigated the functional variability of distinct recombinant UL146 proteins showing that UL146 polymorphisms differentially affect chemokine receptor binding affinity, which could influence HCMV dissemination and pathogenesis (Heo et al., 2015). Hence, it might be speculated that the absence of UL146 gt3, gt5, and gt6 in this cohort is due to a lower virulence compared to the other genotypes. In general, cellular CXC chemokines are divided into two groups depending on the presence or absence of an ELR (Glu-Leu-Arg) motif prior to the CXC motif with an angiogenesis promoting and inhibiting function, respectively. Interestingly, all UL146 genotypes that have been detected in our cohort share this ELR motif but not the missing genotypes gt5 and gt6 (Heo et al., 2008). High levels of ELR chemokines have been suggested to be major mediators of lung disease processes such as bronchiolitis obliterans syndrome (BOS), adult respiratory distress syndrome and pulmonary fibrosis (Belperio et al., 2005)(Keane et al., 2002)(Keane et al., 1997). It can be speculated that increased activity of viral ELR chemokine homologues in the lung could change the balance between angiogenic or angiostatic chemokines in favour of aberrant angiogenesis. This might play a role in why HCMV replication is a risk factor for BOS development (Paraskeva et al., 2011), and might be worth to be addressed in future studies.

A main objective of this study was to identify the relationship between pre-transplant D/R HCMV serostatus and strain diversity and dynamics post-transplantation. The finding that D+R+ patients harbour a higher number of genotypes compared to the other two D/R groups (Figure 3B) indicates that both, donor- and recipient-derived strains contribute to the observed diversity. Although several previous studies found that reinfection with donor-derived strains are more common than reactivation of recipient virus (Grundy et al., 1988)(Chou, 1986)(Sunwen & Norman, 1988), others have argued that infections post-transplantation are approximately equally donor- and recipient-derived (Manuel, Pang, et al., 2009). Reports on the detection of HCMV in tissues of non-immunocompromised individuals (Schonian et al., 1993)(Kytö et al., 2005)(Kraat et al., 1992)(Hendrix et al., 1997)(Meyer-König et al., 1998) further underline the potential of HCMV transmission with the allograft, yet the extent of transmission remained controversial. In the present study, we could demonstrate HCMV-DNA positivity in 5/10 pre-transplant lungs of HCMV-seropositive donors, with up to three HCMV genotypes present in small HCMV-DNA positive tissue sections of a single donor lung. Assuming that the detected HCMV-DNA reflects replicating rather than latent HCMV-DNA, these findings indicate focal points of HCMV reactivation of either a single or multiple HCMV strains. Of note, in the HCMV-positive cases reactivation must have started already before surgical organ removal, which is likely given that all donors were in the intensive care unit and under ventilation for >24 hours. Interestingly, the focal distribution of HCMV-DNA in the human lung resembles the murine CMV infection model in which the authors also observed a ‘patchwork pattern’ of focal reactivation and recurrence (Kurz et al., 1999)(Kurz & Reddehase, 1999). Taken together, our data strongly support the findings that HCMV transmission by the allograft of an HCMV-seropositive donor is very likely, yet either the extent thereof or whether it occurs or not may vary among donors. In our present study 2/15 D+R+ patients analysed longitudinally were solely infected with a single HCMV strain despite an overall higher diversity in this patient group. Nevertheless, our findings point towards the donor’s lung as an important contributor to HCMV strain transmission in both HCMV seropositive and seronegative recipients. Strategies to target this reservoir in pre-transplant lungs of HCMV seropositive donors are promising. Recently, the ‘shock and kill’ approach in which latent cells are targeted and cleared has been applied to donor lungs. Treatment with the immunotoxin F49A-FTP during ex-vivo lung perfusion has been shown to reduce reactivation to 76% compared to 15% in non-treated, but perfused controls (Ribeiro et al., 2022). We could further show that D+ patients display higher replication dynamics of their mixed HCMV strain populations post-transplantation compared to D-patients, also arguing for the donor-derived strains as the likely cause. It can be speculated that sequential reactivation and dissemination of donor-derived strains post-transplantation can ultimately increase the host’s latency reservoir and gradual reseeding of the lung as well as dissemination into other tissues can contribute to the distribution of HCMV strains (Collins-McMillen et al., 2018). Over time, these processes might result in locally and temporally different replication levels of the distinct strains. This will either be observed as an increase in the genotype numbers and/or as a change in predominance. The latter dynamics pattern occurs more frequently in D+R+ patients which may be explained by its generally richer pool of strains. A previous study demonstrating the sequential occurrence of different predominant strains in D+R-patients receiving organs from the same donor support our view of sequential and/or stochastic reactivation of donor strains from the graft (Hasing et al., 2021), as has also been proposed by the murine CMV model (Reddehase et al., 1994). Additionally, it is presumable that initial replication of the major strain triggers a strain-specific immune response which might be well controlled thereafter while the initially less prevalent strain becomes predominant in further episodes due to limited cross-protection (Klein et al., 1999)(Wang et al., 2021). Antiviral medication, in contrast, does not substantially interfere with the replication dynamics since no difference in administration of antivirals was observed between patients with and without dynamics or between the different serostatus groups.

In-depth short amplicon sequencing over polymorphic regions is a highly sensitive strategy to decipher mixed genotype infections but may underestimate the true strain diversity due to the small genomic regions analysed and to the lack of information on linkage patterns. To overcome these limitations, we additionally performed long amplicon haplotype sequencing on a subset of 24 samples of 20 patients of whom 10 patients were mixed infected. The four haplotype target regions (F1, F3, F4, F5) cover about 10-fold larger genomic regions than our short amplicon approach. The advantage of long reads to resolve distinct haplotypes in mixed populations became particularly obvious in patients in whom we found haplotypes with partially shared genotypes (Figure 6 – figure supplement 2). Hence, the lack of information on the arrangement of genotypes on individual genomes by short amplicon genotyping approaches, might help to understand why correlating specific genetic variants to clinical observations may lead to inconsistent results (Wang et al., 2021). In one patient, same genotype assignments were shared between hap1 and hap2 in one locus (UL139) and hap2 and hap3 in other loci (UL144, UL146), suggesting that hap2 had arisen by intra-host recombination between hap1 and hap3. The low number of single nucleotide polymorphisms between the same genotypes, which could have been accumulated over time in the patient, would have supported this conclusion. However, the substantial differences across the full length haplotype sequences makes a recent intra-patient generation of a recombinant haplotype unlikely since within-host haplotype sequences are highly stable on short timescales as shown in our study and by others (Götting et al., 2021)(Dhingra et al., 2021)(Hage et al., 2017)(Cudini et al., 2019). Additionally, almost identical sequences of the respective haplotypes can be found in the database, further pointing towards transmission from another host. Also, identical haplotypes were observed across patients of our cohort and in one patient (LTR-41) we identified a previously described non-recombinant strain, supporting the view that HCMV diversification has occurred early in human history (Suárez, Wilkie, et al., 2019)(Mattick et al., 2004)(Bradley et al., 2008). Both approaches used in our study, short- and long–read sequencing, suggest an upper limit to intra-host HCMV genetic diversity with a maximum of four genotypes and three haplotypes, respectively, in our cohort of lung transplant patients. Although long read sequencing of longitudinal samples is suitable for directly proving within-patient recombination no direct evidence was seen in the analysed genomic regions. Hence, from our data, it appears that the overall within-host HCMV diversity predominantly results from a mixture of genomically distinct strains rather than from newly emerging variants which confirms previous studies (Cudini et al., 2019)(Suárez, Wilkie, et al., 2019)(Suárez et al., 2020).

In conclusion, by HCMV genotype and haplotype determination of clinical samples, we demonstrate the extent to which the donor lung can contribute to HCMV strain diversity and dynamics after transplantation. We suggest that rapid intra-host dynamics of a limited number of HCMV strains might allow quick adaptation to changing environments and less so by enhancing diversity through recombination. Understanding the forces affecting HCMV population diversity and dynamics is an essential step for treatment and vaccine development.

## Materials and Methods

### Study design and sample collection

For this retrospective study, we analysed routine HCMV-DNAemia monitoring data for EP and BAL specimens of LTRs post-transplantation. Between 2016 and end the of 2018, 286 patients received a lung transplant at the Medical University of Vienna. As a standard regimen, all LTRs receive immunosuppressive treatment and antiviral prophylaxis for at least three months post-transplantation. To investigate strain dynamics, we selected for LTRs with 1) at least two active HCMV infection episodes (>10^2^ copies/mL) in either EP or BAL and 2) with at least one sample with >10^3^ copies/mL (to maximise the chance for long range amplicon PCR). We defined positive HCMV-DNAemia sample points as distinct episodes if between two sample points 1) the viral load declined below 10^2^ copies/mL or 2) if there was a time interval of three months. All EP and BAL specimens were stored at −20°C. Of the total 339 specimens from 53 LTRs that were initially selected, 81 specimens were not available. DNA extraction of 67 was unsuccessful, and PCR amplification of 28 samples resulted in no amplicons. Consequently, three LTRs were excluded. Two to 15 samples were collected from each LTR and sampling time points ranged between four and 1476 days post-transplantation. Clinical information about the patients of the final cohort was retrieved from medical records.

### DNA extraction and viral load quantification

DNA was isolated from 250 µL of BAL or EP samples using the QIAamp Viral RNA Mini kit (Qiagen) as described in the manufacturer’s protocol and eluted in 35 µL elution buffer. HCMV DNA was quantified by in-house HCMV-specific qPCR as previously described (Kalser et al., 2017) and stored at 4°C or −20°C for further steps.

### Short and long-range amplicon PCR

Short-range amplicon PCRs for the five polymorphic regions gN, gO, UL6, UL139 and UL146 of HCMV-DNA positive samples were performed as described previously (Brait et al., 2022), but with minor modifications. To save limited DNA material, UL6 and the first step of the nested UL146 PCR was multiplexed. Also, gO PCRs, consisting of three primers, were multiplexed. For samples with low viral loads (<10^3^ copies/mL) and sufficient material availability, the usual DNA template amount of 5 µL was doubled. The viral load sensitivity of our long-range PCR was tested for BAL and EP samples (data not shown). Based on that, BAL samples with >5×10^3^ copies/mL and EP samples with > 5×10^4^ copies/mL were initially amplified with long range PCR. In addition to previously described amplicons F3 and F4 (Brait et al., 2022), F1 and F5 amplicons were designed and tested to cover more polymorphic regions with long reads. F1, F3 and F4 PCR primers were multiplexed. PCR of samples with lower viral loads and EP samples, in general, were performed with the doubled template amount of 20 µL, if available. Samples with unsuccessful long-range PCR were genotyped using short range PCR. All primers are depicted in Figure 1, and Supplementary Table 6 lists detailed descriptions of PCR primers. PCR product lengths were confirmed on analytical agarose gels and concentrations quantified by Qubit prior to Illumina or PacBio library preparation.

### Illumina and PacBio sequencing

Library preparation and sequencing for Illumina sequencing were performed as described previously with a few adaptations (Brait et al., 2022). Briefly, amplicons were pooled equimolarly and 2 ng input DNA was used to generate a 4 nM library using the Nextera XT library preparation and index kit, followed by paired-end sequencing (150 cycles, v2 and v2 Micro kits) on an Illumina MiSeq with automatic adapter trimming. Fastq raw reads that passed filters were imported into CLC Genomics Workbench 21.0 (Qiagen) for further analysis.

According to manufacturer’s protocol, pooled long range amplicons were purified using the Qiaquick Spin Kit and eluted in 50 µL elution buffer. After quantification and purity determination with NanoDrop 1000 tool (Peqlab) and Qubit 2.0 fluorometer (Thermo Fisher), samples were submitted to the Next Generation Sequencing Facility at Vienna BioCenter Core Facilities for SMRT bell Amplicon library preparation. Sequencing was performed on a PacBio-Sequel system for 20 hours. Twenty-four samples were multiplex sequenced on two lanes.

### Bioinformatical workflows for PacBio and Illumina reads

#### PacBio CCS generation

CCS reads were generated from PacBio raw reads using the *ccs* tool (https://github.com/PacificBiosciences/ccs) with a minimum predicted read quality of 0.99 (>99% accuracy) and a minimum of three full passes per strand. To avoid CCS of potential PCR-mediated heteroduplexes of different strains, the *--by-strand* option was used whereby CCS for forward and reverse strands are generated and analysed separately. Next, CCS were demultiplexed using *lima* (https://github.com/pacificbiosciences/barcoding/) and resulting bam files imported into CLC Genomics Workbench 21.0.3 (Qiagen) for further analysis.

#### HCMV haplotype determination

For each of the four long-range amplicon regions, we compiled reference databases to be used for read mapping. First, we extracted the corresponding amplicon region from 236 publicly available HCMV whole-genome sequences and aligned them using MUSCLE 3.8.31. Pairwise distances were calculated in MEGA X 10.2.4, and only sequences with >150 nucleotide differences in the respective region were filtered for the final reference databases. These resulted in 33, 23, 16 and 93 reference sequences for F5, F1, F3 and F4, respectively (Supplementary Table 7a). Trees were generated in MEGA X 10.2.4 using the UPGMA method and displayed with Evolview v3 (Subramanian et al., 2019) (Figure 5a).

Firstly, human reads were excluded, and reads were length trimmed according to the amplicon length (F1 7.5-7.8 kb; F4 6.45-6.75 kb; F1 7.8-8.1 kb, F5 6.2-6.5 kb). Using the Long Read Support plugin for CLC Genomics (Qiagen), reads were mapped against the filtered reference sequences with default settings and extractions of the consensus haplotype sequences were generated (low coverage threshold: 3, ambiguity code N insertion with the noise threshold set to 0.3 and the minimum nucleotide count to 3). As a control step, all initial, length trimmed reads were re-mapped to the new consensus haplotype(s), and the new consensus sequence was extracted. All final haplotypes of each sample were aligned and checked for their uniqueness. Potential chimeras were inspected visually by checking if final haplotypes could be reconstructed by combining segments of two more abundant haplotypes. Lastly, all unmapped reads of the first mapping of each amplicon were again mapped to all final haplotypes to confirm that all unmapped reads for a certain amplicon belong to any of the other three amplicons.

#### HCMV genotype determination

Q30 quality trimmed Illumina fastq reads were filtered for reads >80 bp in length and human genomic reads were excluded. Default mapping parameters were used for match/mismatch scores, insertion/deletion costs, and a length fraction of 0.3 and a similarity fraction of 0.95. Only mappings with >10 reads and a consensus length of at least 75% of the reference sequence were chosen to extract consensus sequences. A noise threshold of 0.3 and a minimum nucleotide count of 3 was applied to insert ambiguity codes (N). All resulting genotype consensus sequences were aligned, screened visually and unique sequences counted as genotypes. PacBio-derived haplotypes were genotyped using blastn (match/mismatch= 2/-3, gap costs = existence 5, extension 2) and a self-assembled BLAST database (Supplementary Table 7b).

For genotyping, five highly polymorphic regions of UL6 (127-388), gO (691-987), gN (1-379), UL139 (187077-186395, CDS: 1-414) and UL146 (181341-180571, CDS: 1-360) were used based on sequences provided previously (Suárez, Musonda, et al., 2019). In total, 47 reference sequences were used. For long amplicon reads, genotyping with 34 additional references was performed for regions gH (1-177), UL144 (1-500), UL9 (57-430), UL10 (0-762), UL11 (194-623) and gB (1138-1619). All listed nucleotide positions are in reference to strain Merlin (GenBank: AY446894.2). Reference sequences for genotyping are provided in Supplementary Table 7b.

### Donor lung tissue processing

Lung tissues were removed for pre-transplantation size adjustment and would have been discarded otherwise. Middle lobe sections of 1-2 cm^3^ of ten HCMV-seropositive donors were collected in phosphate-buffered saline (PBS) and stored at 4°C for processing on the same day (solution termed supernatant, short SN). Bigger sections were divided, and a part submerged in RNAlater™ solution (Invitrogen) for storage at −20°C. Next, sections were minced with sterile single-use scalpels, weighted in a 50 mL falcon tube, and digested with Hanks’ balanced salt solution supplemented with 0.15% Collagenase D (Roche) for 60 min at 37°C on an orbital shaker at 300 rpm. The tissue solution was diluted with PBS, vortexed vigorously and strained through a 70 µm cell strainer to obtain a single cell suspension. After centrifugation (500 rpm, 10 min, 4°C), red blood cells were lysed with RBC solution (Invitrogen™ eBioscience™) and the remaining cells were counted using an automated cell counter (Nexcelom). Dead cells were excluded using trypan blue. Both, 1 mL of SN and the single cell suspension (at most 10^6^ cells) were separately transferred into 2 mL of lysis buffer and eluted in 50 μL elution buffer using the bead based NucliSens EasyMagextractor (BioMérieux). HCMV-specific qPCR positive samples were genotyped using short-range amplicon PCRs and the NGS workflow described earlier.

### Statistical Analysis

Statistical calculations were performed in GraphPad Prism version 9.0.0. *P* < 0.05 was considered statistically significant in all tests.

### Data availability

Raw sequence data have been deposited in NCBI Sequence Read Archive under BioProject ID PRJNA803978. Haplotype sequences generated in this study and with identities < 98% to publicly available sequences were submitted to GenBank with the accession no. OM835733-OM835736 (Supplementary Table 4b).

## Data Availability

All data produced in the present work are contained in the manuscript.

## Acknowledgments

We acknowledge the support and technical assistance of Barbara Jilka, Andreas Rohorzka, Sylvia Malik, Michaela Binder, Barbara Dalmatiner and Gabriele Sigmund. Lastly, we want to thank Sylvia Knapp for her valuable advice.

## Additional information

### Funding

This work was funded by a grant from the Austrian Science Fund (FWF) to Irene Goerzer (project number: P31503-B26).

### Declarations

This study was approved by the Ethics Committee of the Medical University of Vienna under EK-number 1321/2017. All data were pseudonymised before analyses.

### Author contributions

## Additional files

### Supplementary files

Supplementary Tables 1-7b, Supplementary_Tables_28022022.xlsx

Transparent reporting form, transparent_reporting_28022022.pdf

## Competing Interests

No competing interests declared.

## Notes

### Competing Interest Statement

The authors have declared no competing interest.

### Funding Statement

This study was funded by the Austrian Science Fund (FWF) project number: P31503-B26.

### Author Declarations

This study was approved by the Ethics Committee of the Medical University of Vienna under EK-number 1321/2017. All data were pseudonymised before analyses. Informed patient consent was not required.

